# Association of Graph-based Spatial Features with Overall Survival Status of Glioblastoma Patients

**DOI:** 10.1101/2022.12.16.22283587

**Authors:** Joonsang Lee, Shivali Narang, Juan Martinez, Ganesh Rao, Arvind Rao

## Abstract

**Background and purpose:** Glioblastoma multiforme (GBM) is the most common malignant brain tumor with less than 15 months median survival. To aid prognosis, there is a need for decision tools that leverage diagnostic modalities such as MRI to inform survival. In this study, we examine higher-order spatial proximity characteristics from habitats and propose two graph-based methods (minimum spanning tree and graph run-length matrix) to characterize spatial heterogeneity over tumor MRI-derived intensity habitats and assess their relationships with overall survival as well as immune signature status of patients with GBM.

**Material and methods:** A data set of 74 patients was studied based on the availability of post-contrast T1-weighted and T2-weighted fluid attenuated inversion recovery (FLAIR) image data in The Cancer Image Archive (TCIA). We assessed the predictive value of MST- and GRLM-derived features from 2D images for prediction of 12-month survival status and immune signature status of patients with GBM via a receiver operating characteristic curve analysis.

**Results:** For 12-month survival prediction using MST-based method, sensitivity and specificity were 0.82 and 0.79 respectively. For GRLM-based method, sensitivity and specificity were 0.73 and 0.77 respectively. For immune status, sensitivity and specificity were 0.91 and 0.69, respectively, for the GRLM-based method with an immune effector.

**Conclusion:** Our results show that the proposed MST- and GRLM-derived features are predictive of 12-month survival status as well as the immune signature status of patients with GBM. To our knowledge, this is the first application of MST- and GRLM-based proximity analyses for the study of radiologically-defined tumor habitats in GBM.

## Introduction

Glioblastoma multiforme (GBM) is a common primary brain tumor known for its aggressive malignant behavior. Prognosis for patients with GBM remains very poor with the median overall survival duration between 12 months and 15 months despite multimodality treatments such as surgical resection followed by combination of radiation therapy and chemotherapy (temozolomide) ^1,2^. Several studies have been proposed to improve the diagnostic performance of MRI for cancer using various techniques such as computer-based image analyses ^3,4^, imaging features analysis ^5-7^, machine learning techniques ^8^, imaging-genomics analysis ^9,10^.

From MRI analysis studies of GBM patients, it has been suggested that intensity-level heterogeneity within the tumor is indicative of multiple tumor regions with distinct MRI intensity characteristics that might respond differently to treatment regimens ^11^. This has implications for the assessment of patient prognosis in GBM ^12,13^. Based on multiparametric measurements of different MRI sequences, these habitats characterize regional variations in blood flow, cell density, and necrosis ^11^. Apart from the abundance of these habitats, the spatial extents and proximity of these habitats have physiologic and clinical relevance for assessment of treatment response ^11,14^. In this study, we identified four distinct groups of pixel intensities (habitats) within the tumor ROI across different MR sequences and characterized the spatial relationships of these derived habitats with graph-based methods such as a minimum spanning tree (MST) construction and graph run-length matrices (GRLM). Previously, graph (MST-derived) features have been used to understand the proximity relationships between distinct immunohistochemical entities (cell types) within hematoxylin and eosin (H&E) pathology slides ^15^. These MST-based features successfully distinguished samples of high and low lymphocytic infiltration extent with a classification accuracy greater than 90% ^15^. Based on the success of this approach, we hypothesized that characterizing the spatial relationship between radiologically-distinct habitats of a tumor using MST or GRLM approaches might have predictive value for underlying clinical outcome in GBM.

Another graph-based characterization called GRLM ^16^ was also used to characterize the spatial heterogeneity of a tumor to predict clinical outcome in GBM, based on the idea of graylevel run-length matrices. The gray-level run-length method is one of popular methods extracting high order statistical features in texture analysis ^17^. In this study, we used GRLM to compute the runs of radiologically defined tumor habitats on a region of interest (ROI) image to obtain a runlength matrix instead of counting runs of pixel intensities.

The purpose of this study is to evaluate the prognostic significance of higher-order spatial proximity characteristics from habitats using quantitative metrics derived from graph based methods. We aim to investigate the association of MST and GRLM-derived spatial proximity features of these tumor habitats with overall survival of GBM patients as well as predicting immune signature status. In the broader context, this study aims to investigate relationships between imaging and phenotypic characterizations of the tumor, thereby augmenting the foundation for population-based correlation studies in GBM.

## Materials and methods

### Data

A data set of 74 patients was studied based on the availability of post-contrast T1-weighted and T2-weighted fluid attenuated inversion recovery (FLAIR) image data in The Cancer Genome Atlas (TCGA), The Cancer Imaging Archive (TCIA - http://www.cancerimagingarchive.net/). The dataset consisted of 25 female and 49 male patients with de-novo (primary) GBM. The patient demographics are summarized in Table 1. The mRNA expression data and clinical data such as survival information for these cases ^18^ were obtained from the cBioPortal for Cancer Genomics (http://www.cbioportal.org). In this study, the previously defined immune effector and immune suppressor response ^19^ were used to derive the immune gene signature status using single-sample gene set enrichment analysis: ssGSEA for each patient ^20-23^. For predicting immune signature status, 34 patients were used based on availability among 74 patients. The MR images were preprocessed with registration, non-uniformity correction using N3 ^24^, pixel reslicing, and intensity normalization ^25^ before subsequent analysis. Registration of the T1 post-contrast image and T2 FLAIR image along with non-uniformity correction for MRI-artifacts were performed using the Medical Image Processing, Analysis, and Visualization (MIPAV) software ^26^. The FLAIR MR image is registered to the T1 post-contrast image using affine transformation with 12 degrees of freedom along with trilinear interpolation. Segmentation was a semi-automated process for which MITK3M3 Image analysis toolkit was used. The clinicians used this tool to contour the tumor region on multiple slices with interpolation performed to obtain a 3D volumetric tumor mask. This step was performed independently on T1-post contrast as well as T2-FLAIR. In all the processes, readers were blinded to clinical/molecular characteristics. Pixel reslicing was performed using the NIFTI toolbox in MATLAB to make pixel sizes isotropic (1mm). The resulting T1 post-contrast image and T2 FLAIR image after preprocessing is shown in Figure 1.

**Table 1.** Patient demographics

| Characteristics | OS ≤12 mo. | OS >12 mo. | Total |
| --- | --- | --- | --- |
| Male (n) | 19 | 30 | 49 |
| Female (n) | 12 | 13 | 25 |
| Age (y) | 61.6 ± 15.3 | 54.5 ± 15.4 | 57.5 ± 15.7 |
| Overall survival (mo.) | 5.9 ± 3.0 | 24.8 ± 12.7 | 16.9 ± 13.6 |

The data for age and overall survival are means with standard deviations; OS – Overall survival

**Figure 1.**
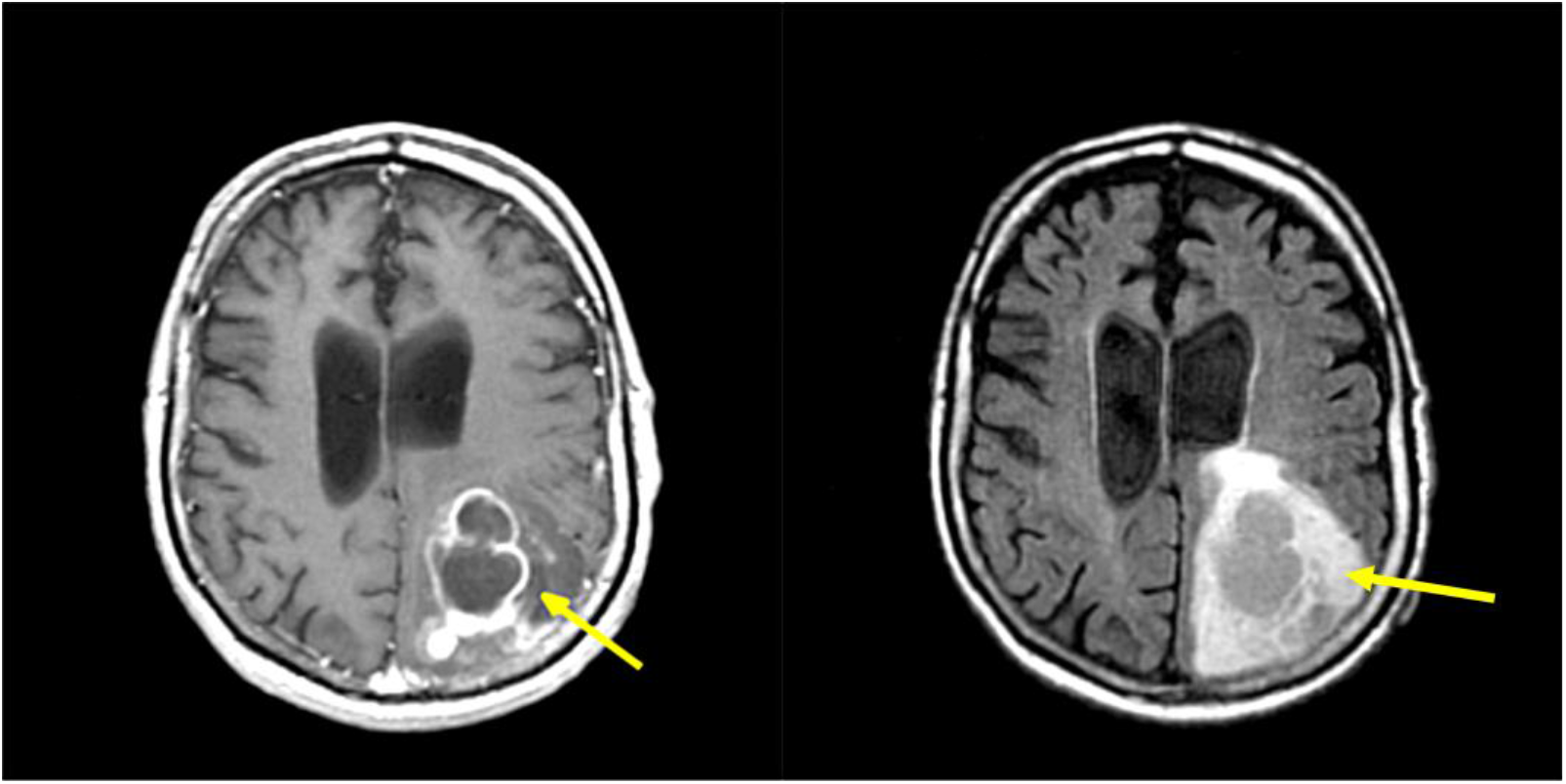
T1 post-contrast image (left) and T2 FLAIR image (right) after preprocessing. Arrows point to the enhanced tumor areas.

### Region of interest (ROI) Delineation

A tumor ROI was segmented semi-automatically by two experts (radiologists) using the Medical Imaging Interaction Toolkit (MITK). The extent of tumor was defined using the contrast enhancing tumor region within the T1-post contrast image, and the areas of solid tumor, infiltrating tumor and edema regions within the T2-FLAIR image. The slice with the maximum tumor area in T1 post-contrast image and the corresponding slice from the T2 FLAIR image were selected for both MST and GRLM analysis. The pixel intensity values within the ROI for each patient were scaled to lie between zero and one by linear transformation, and then fitted using a two-component Gaussian mixture model (GMM) ^27,28^. The threshold between two Gaussian groups was determined by calculating the average of the means of the two Gaussian populations underlying the low intensity pixel group and high intensity pixel group within the tumor, following a process similar to prior work ^28^.

### Minimum spanning tree (MST) and graph run-length matrices (GRLM)

A spanning tree of a graph represents a tree that connects all the vertices. Each edge has an associated weight (or length), the weight of a tree represents the sum of all weights of its edges. Then, the MST can be defined as a spanning tree with the minimum weight of a tree among any other spanning trees.

The gray-level run-length matrix *M*(*i, j*) is a popular method in texture analysis that measures the variation of the pixel intensities to quantify intuitive qualities such as smoothness, coarseness, and roughness. Gray-level run-length can be defined as the number of runs with pixels of gray level *i* and run length *j* in a given direction. Basically, run-length matrix provides the coarseness of a texture in a specified direction. Runs of data represent sequences in which the same data value, gray level intensity, occurs in many consecutive pixel elements. In general, fine texture or high frequency tends to have more short runs with similar gray level intensities and coarse texture or low frequency tends to have longer runs of similar gray level intensities.

### Feature extraction with minimum spanning tree (MST)

In this study, we used two different MR sequence images (T1 post-contrast image and T2 FLAIR image). For each MRI image-sequence, the pixels within the tumor ROIs was separated into low and high intensity group (habitat) using the two-component GMM. Four binary masks were prepared from these groups (for every pairwise combination of habitats). A two-dimensional grid line was overlaid on each binary mask. The grid lines were equally spaced with a distance of 8 pixels (8mm × 8mm), chosen empirically. Next, we computed the coordinate of the centroid that specifies the center of mass of the region from each habitat inside of the small bounding grid box ^29^. Coordinates from all grid boxes in each habitat were combined into one map. An MST was constructed across all these co-ordinates as vertices. Thus we have four MSTs (one for each habitat: T1 high intensity group, T1 low, T2 high, or T2 low intensity group, respectively). Figure 2 shows MST of each of the four groups on an ROI map. The mean, median, standard deviation, skewness, kurtosis, min/max ratio, and disorder of the branch lengths in MST were computed to obtain a set of seven features ^15^ (for each habitat. Expressions for the features are listed below: Mean edge weight, *f*_*µ*_:

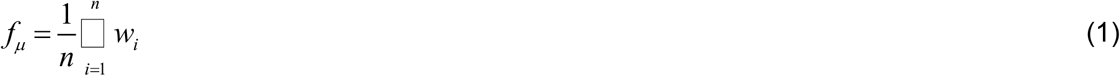

where *w*_*i*_ is an individual weight or branch length in MST.

**Figure 2.**
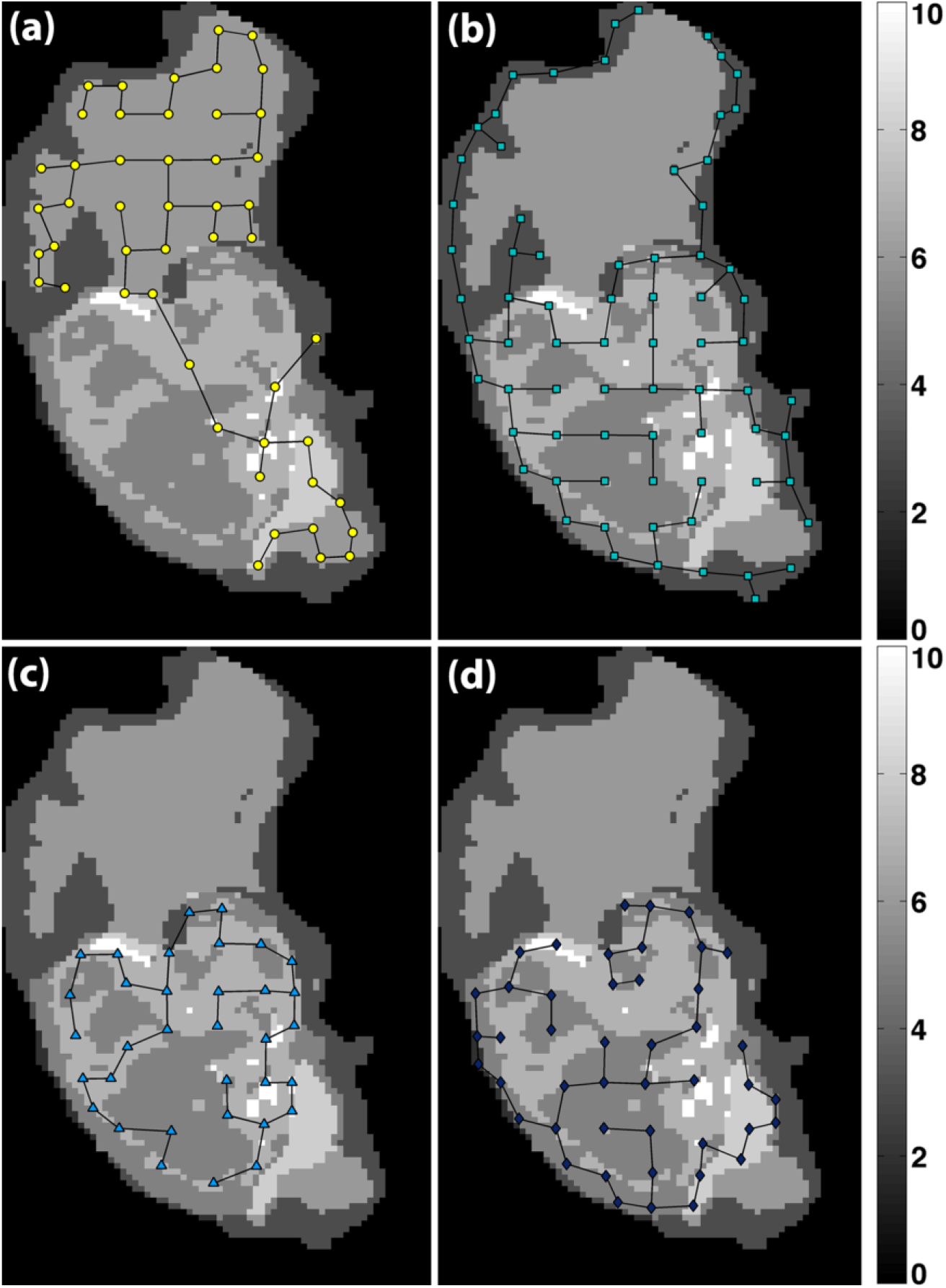
MSTs of the four habitats within the tumor ROI (a) T1 post-contrast high pixel intensity group, (b) T1 post-contrast low pixel intensity group, (c) T2 FLAIR high intensity pixel group, and (d) T2 FLAIR low intensity pixel group, respectively. Different gray levels within the ROI represent different habitats and their overlapping areas.

Median edge weight, *f*_*median*_, is the value separating the higher half of branch lengths from the lower half.

Standard deviation of the distribution of edge weights, *f*_*α*_ :

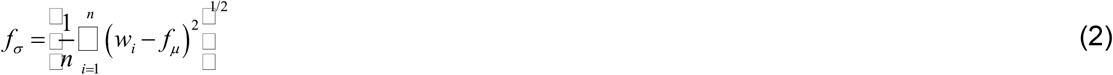

Skewness of distribution of edge weights, *f*_*skewness*_:

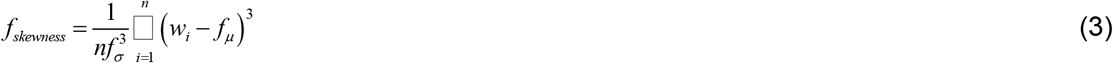

Kurtosis of distribution of edge weights, *f*_*kurtosis*_,:

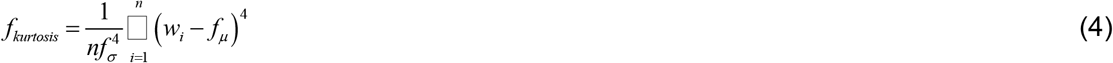

Finally, the min/max ratio, *f*_*r*_, is the ratio between maximum of **W** divided by the minimum value of **W** where **W** is a set of weights (branch lengths), **W** ={*w*_1_,*w*_2_, …, *w*_*n*_} .’Disorder’ is the standard deviation *f*_*α*_ divided by the mean value of **W**, *f*_*μ*_.

### Feature extraction with graph run-length matrices (GRLM)

Similar to the construction of the MST, we computed the coordinate of the centroid from each habitat inside of the rectangular grid box. For the extraction of GRLM features, we combined all coordinates from all four habitats into one set (map). Again, these four habitat groups represent T1 high- and low-intensity pixel groups and T2 high- and low-intensity pixel groups. Figure 3 shows an ROI map with all coordinates (across all four habitats) as vertices. Then, we constructed a Delaunay triangulation ^30^ to connect these vertices. There are several ways to triangulate any given set of points and a Delaunay triangulation is one of the most widely used in scientific computing of various applications. In the Delaunay triangulation, all triangles for a set of points will have empty circumscribed circles ^30^.

**Figure 3.**
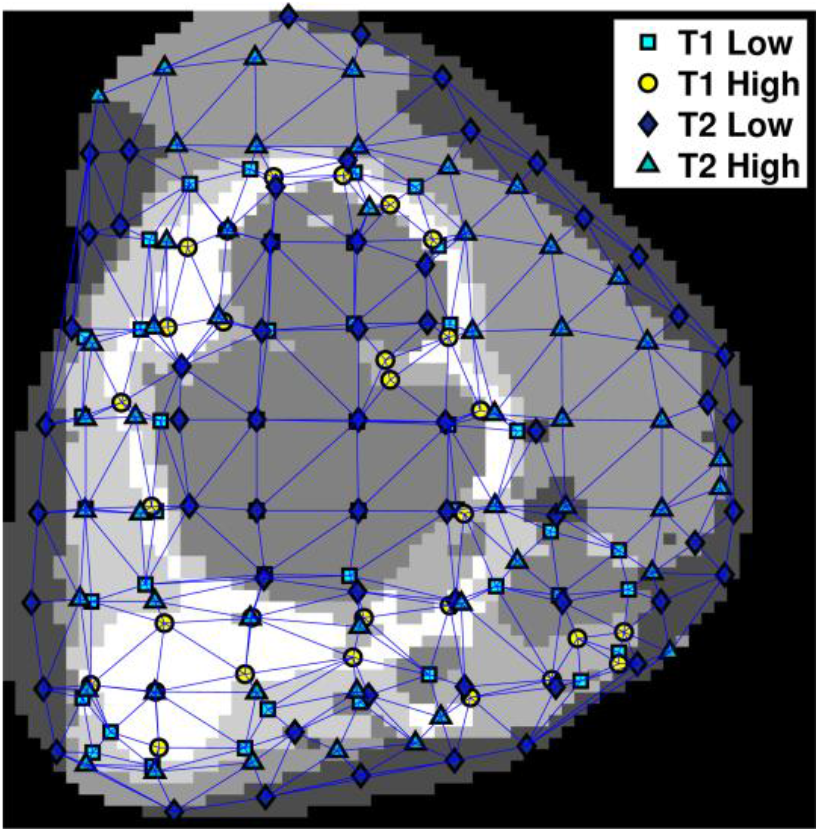
An example of ROI map with a Delaunay triangulation across all vertices. Different gray levels within the ROI represent different habitats and their overlapping areas.

After constructing the ROI map with the Delaunay triangulation, a run-length matrix was computed. In this study, we used the GRLM method in the manner proposed by Tosun et al. for histopathological image segmentation ^16^ because this aims to represent the spatial separation between point set entities. GRLM *G*(*t, l*) can be defined as the number of graph-edge runs with an edge type *t* and a path length *l* for a single node. The algorithm starts from the initial node to the furthermost node in the path within a circular window. Figure 4 shows the calculation of a graph run-length matrix for a single node. For the entire region, the algorithm accumulates the run-length matrices of the nodes in an ROI.

**Figure 4.**
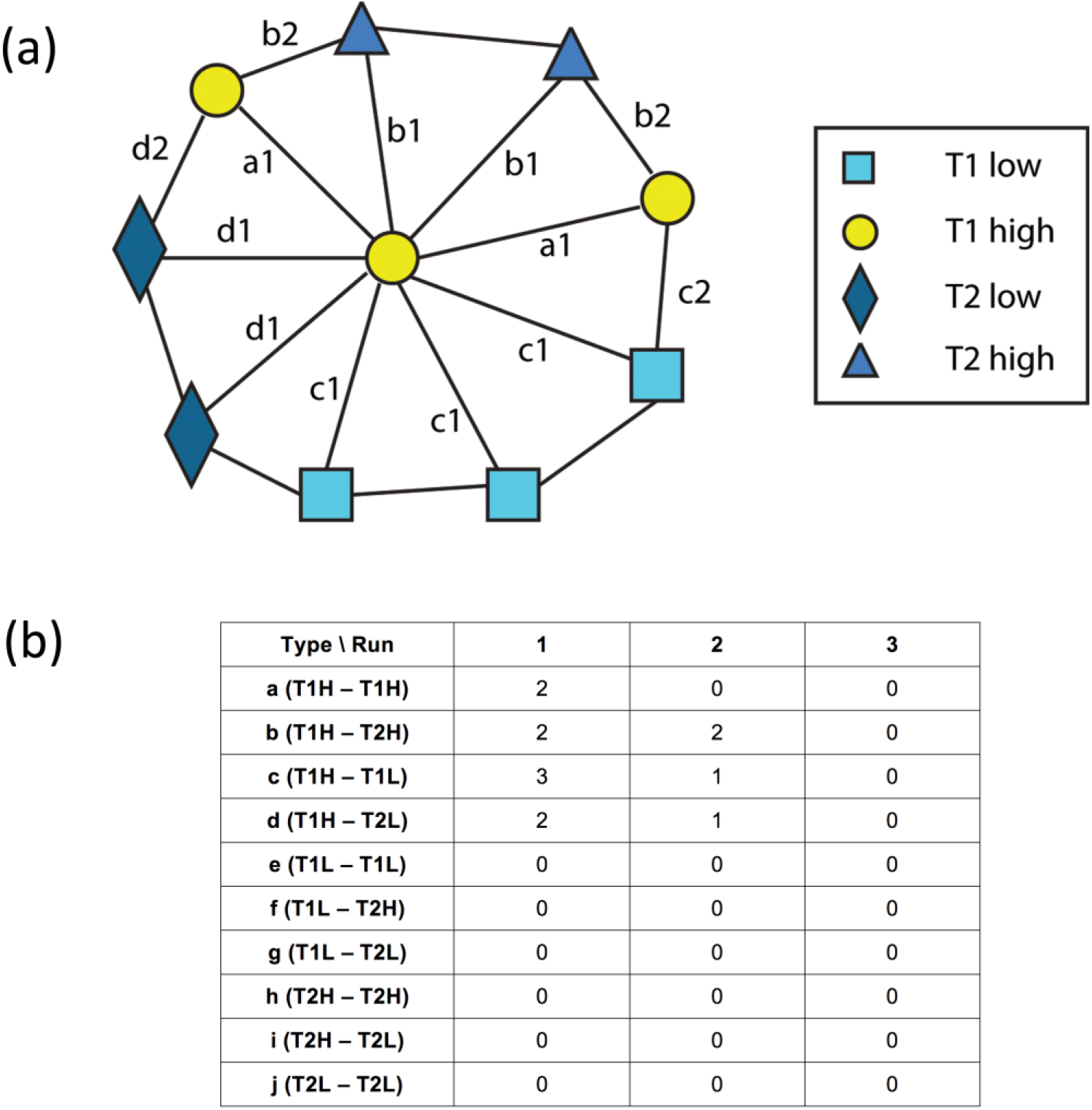
(a) Illustration of a single initial node located at the center and (b) a graph run-length matrix for this single initial node. This method was adapted from Tosun et al.

For extracting GRLM-based features, we used six measurements: short path emphasis (SPE), long path emphasis (LPE), edge type nonuniformity (ETN), and path length nonuniformity (PLN). The expressions for these features are listed below:

Short path emphasis (SPE) and SPE(*t*) for each edge type *t* :

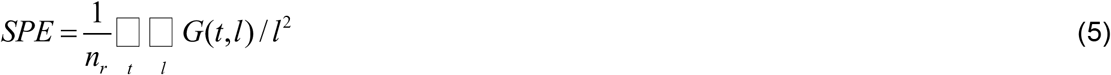

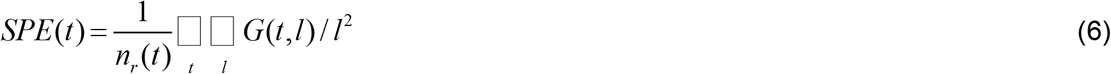

where *n*_*r*_ is the total number of runs in the GRLM and *n*_*r*_ (*t*) is the total number of runs corresponding to edge type *t*.

Long path emphasis (LPE) and LPE(*t*) for each edge type *t*:

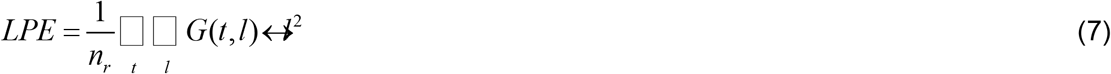

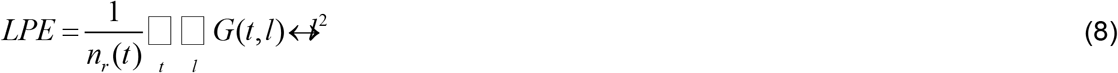

Edge type nonuniformity (ETN) and path length nonuniformity (PLN):

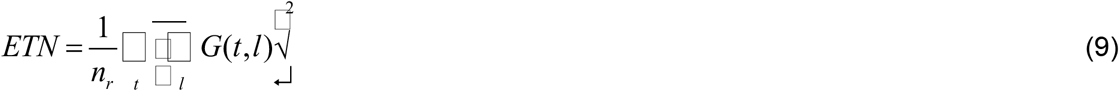

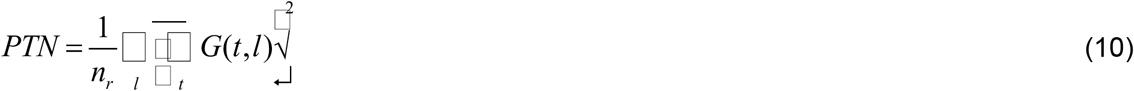

### Classification of Immune signature status

The immune signature scores from the TCGA cohort were dichotomized at the median value; either an up regulated as signature score > median value or down regulated as signature score ≤ median value. This binary designation is used as the class label in the classification task. These gene signatures associated with immune status in GBM, immune effector response and immune suppression response, have been previously validated within GBM and were evaluated in every patient in our dataset ^19^.

### Statistical analysis

A total of 28 MST based features (seven MST-features from each of the four habitats) and 24 GLCM based (SPE, LPE, ETN, PTN, and 10 SPE(*t*) and 10 LPE(*t*)) features for 10 edge types were extracted and analyzed in this study. Survival was dichotomized at the 12 month time point (i.e. ≤ or > 12 months) based on the median overall survival duration ^1,2^ and imbalance in sample size of the two survival groups was controlled using class-proportional sampling ^31^ in Waikato Environment for Knowledge Analysis (WEKA v3.7.12) ^32^. Classification of the survival labels using all of the MST features was performed using random forest (RF) classification ^33^ (10000 trees using random forest classifier within WEKA). This performance was evaluated using the receiver operating characteristic (ROC) curve. The ranking of the features was also obtained using a classifier-based attribute evaluator (where RF was set as the classifier) within WEKA. Sample size imbalance between classes is handled using class-proportional sampling ^31^.

## Results

Classifier models were obtained using the random forest classification approach ^33^ with 5-fold cross-validation for the prediction of 12-month overall survival status based on all of the features derived from MST and GRLM-based methods, respectively. Figure 5 shows the ROC curves for the classifiers for MST and GRLM. The optimal cutoff point was determined by maximizing the sum of sensitivity and specificity. The results for the area under the ROC curves, the true positive rate, and true negative rate were summarized in Table 2. The area under the ROC curves was 0.832 for MST and 0.773 for GRLM. The accuracy was 0.807 for MST and 0.747 for GRLM computed using Eq. (11)

**Table 2.**
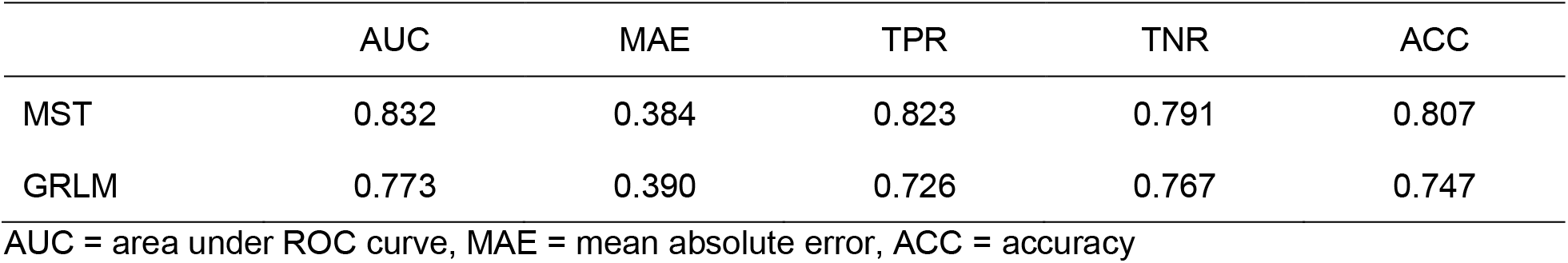
The results of the ROC analysis for the 12-month survival prediction

|  | AUC | MAE | TPR | TNR | ACC |
| --- | --- | --- | --- | --- | --- |
| MST | 0.832 | 0.384 | 0.823 | 0.791 | 0.807 |
| GRLM | 0.773 | 0.390 | 0.726 | 0.767 | 0.747 |

AUC = area under ROC curve, MAE = mean absolute error, ACC = accuracy

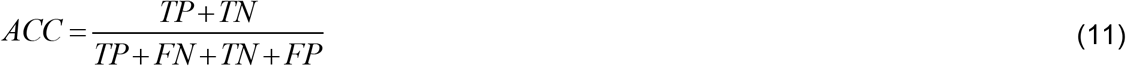

where TP, FP, TN, and FN represent true positive, false positive, true negative, and false negative, respectively. The most important features based on rank (top five) are ratio of T1-low MST, ratio of T2-high MST, ratio of T1-high MST, disorder of T1-low MST, and standard deviation of T1-high for MST and LPE1, SPE4, LPE4, LPE10, and SPE10 for GRLM.

**Figure 5.**
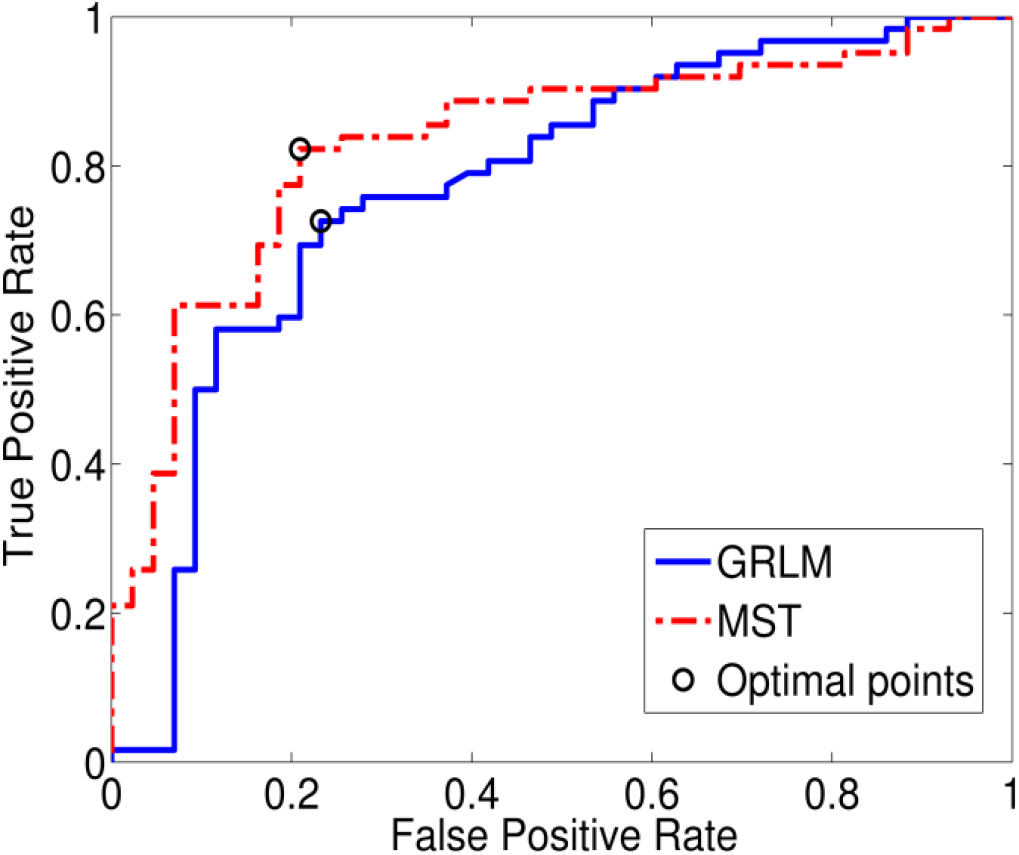
ROC curve for prediction of 12-month survival status. The x-axis is the false positive rate (or 1 – specificity); the y-axis is the true positive rate (or sensitivity). The area under the ROC curve is 0.832 for MST and 0.773 for GRLM. The optimal cut off points are (0.23, 0.73) and (0.21, 0.82) for GRLM and MST, respectively.

For classification of immune signature status, table 3 summarized the true positive rate, and true negative rate, and classification error for MST and GRLM, and Figure 6 shows the ROC curves for the classifiers for MST and GRLM, respectively. The optimal cutoff points were determined by maximizing the sum of sensitivity and specificity.

**Table 3.** The results of the ROC analysis for the immune status

|  | Immune status | AUC | TPR | TNR | ACC | 95% CI (%) |  |
| --- | --- | --- | --- | --- | --- | --- | --- |
|  |  |  |  |  |  | Sensitivity | Specificity |
| MST | IE | 0.747 | 0.861 | 0.579 | 0.720 | 83.8 – 88.2 | 54.8 – 61.0 |
|  | IS | 0.743 | 0.952 | 0.594 | 0.773 | 93.7 – 96.4 | 56.3 – 62.5 |
| GRLM | IE | 0.681 | 0.694 | 0.632 | 0.663 | 66.4 – 72.3 | 60.1 – 66.2 |
|  | IS | 0.789 | 0.905 | 0.688 | 0.800 | 88.5 – 92.3 | 65.8 – 71.7 |

**Figure 6.**
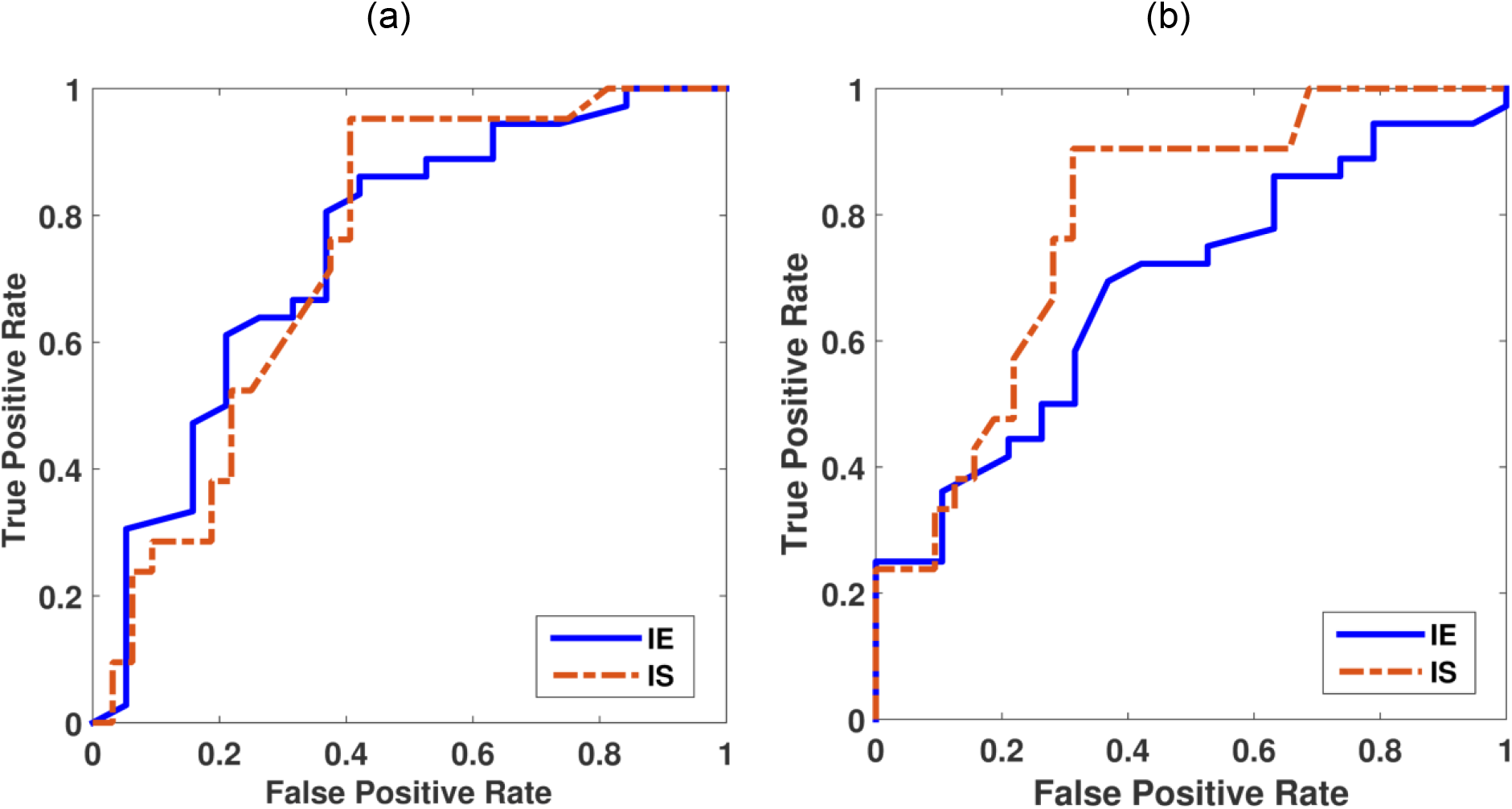
ROC curve for prediction of immune signature status. The x-axis is the false positive rate (or 1 – specificity); the y-axis is the true positive rate (or sensitivity). (a) The area under the ROC curves for MST are 0.747 and 0.743 for IE and IS, respectively, and (b) the area under the ROC curves for GRLM are 0.681 and 0.789, for IE and IS, respectively.

## Discussion

In this study, we identified four distinct groups of pixel intensities (habitats) within the tumor ROI obtained from different MR sequences and separated them into high and low intensities using Gaussian mixture model. All studies were performed in a 2D slice with the maximum tumor area in both T1 post-contrast image and the T2 FLAIR image. These four habitats are considered as distinct entities within an ROI and their spatial relationships are characterized using MST and GRLM approaches. The MST is one of the most common characterization of the spatial proximity of points distributed topologically in space ^34^. Gray-level run-length matrix is also a widely used method in texture analysis that characterizes image texture based on gray-levels run-length of image, introduced by Galloway ^17^.

In this work, we applied these two methods to radiologically defined regions in GBM tumors and investigated the association between graph-based features of habitat proximity from each method (MST or GRLM) with 12-month overall survival status in GBM patients. In this study, we used random forest classification for the following reasons; (i) this classifier can handle large number of features, (ii) it works efficiently in scenarios where the number of instances is smaller than the number of features that are being used for classification, (iii) it is capable of performing cross-validation intrinsically, and (iv) it gives estimates of which variables are important in the classification ^33^.

The proposed MST and GRLM features with existing methods/features could assist towards the assessment of overall survival and serve as a prognostic tool based on routine MRI scans obtained in these patients. To our knowledge, this is the first instance of the investigation of both of these habitat proximity characterizations (MST- and GRLM-based analyses) in the context of multiparametric MRI data and for its application to survival prognostication in GBM. In this study, we used graph-based methods that could have potential advantages to connect between radiologic imaging and cellular evolution within tumors ^28^. These results point to clinically relevant relationships of tumor-derived phenotype with overall survival. Such data could enable generation of valuable hypotheses for the investigation of phenotype relationships based on public domain datasets like TCGA. Further evaluation on clinically-matched patient cohorts with standardized imaging protocols is essential to strengthen evidence for the clinical translation of this finding. In this study, we also showed that these graph-based features are associated with immune signature status in GBM patients with analysis of ROC curves, especially for immune suppressor in GRLM with an AUC value of 0.789.

One potential limitation is that this is a retrospective analysis performed using a publicly available database encompassing various scanning protocols and MRI systems resulting in differences in pixel resolution (256×256 or 512×512), slice thickness (1.4 ∼ 5.0mm), repetition time (4.9 ∼3285.6ms for T1 and 400 ∼ 1100ms for T2), and echo time (2.1 ∼ 20ms for T1 and 14 ∼ 155ms for T2). In this study, we performed image preprocessing steps such as pixel reslicing and intensity normalization to make the MR image aspects comparable across various patients. However, these variations in MR images need to be examined more systematically with both MST- and GRLM-based features.

In this study, we presented graph-based methods for characterizing the spatial proximity of radiologically-defined habitats in GBM tumors, using a minimal spanning tree and graph run-length matrix construction. According to our results, the features from both MST- and GRLM-based methods provided quantitative metrics of image heterogeneity that have prognostic value for patient survival with high accuracy for both methods. We surmise that the spatial proximity features of habitats based on the MST and GRLM approaches may offer a promising method as a clinical prognostic tool in glioblastomas. However, this approach needs to be validated further in an independent patient cohort to confirm its predictive potential.

## Data Availability

All data produced in the present study are available upon reasonable request to the authors

## Acknowledgements

We would like to thank scientific editors, Markeda Wade, Tamara Locke, and Arthur Gelmis, for their help with manuscript editing and suggestions. The authors acknowledge the support of NCI P30 CA016672.

## Data Availability

The datasets generated during and/or analyzed during the current study are available from the corresponding author on reasonable request.

## Author Contributions

Project conception and design were by J.L. and A.R. The data collection and preprocessing were performed by J.L., S.N., J.M and G.R. The software programming, statistical analysis, and interpretation were performed by J.L. The manuscript was written by J.L. and all authors reviewed the manuscript.

